# Testing the effects of a text message intervention on depression and distress among Latino dementia caregivers: a randomized controlled trial protocol

**DOI:** 10.1101/2025.09.16.25335909

**Authors:** Jaime Perales-Puchalt, Mariana Ramírez-Mantilla, Henry P. Moore, Idaly Velez-Uribe, Vanessa Sepulveda-Rivera, Rachel Ruiz, Mónica Fracachán-Cabrera, Yesenia Herrera, Christina Baker, Antonio Miras-Neira, Becky Bothwell, Heidi Anderson, Tina Lewandowski, Francisco J. Diaz, K. Allen Greiner, Kristine Williams, Eric D. Vidoni, Edward Ellerbeck, Jeffrey M. Burns

**Author notes:** **Corresponding Author: Jaime Perales-Puchalt, PhD, MPH,** KU Alzheimer’s Disease Research Center 4350 Shawnee Mission Parkway, Fairway, KS 66205.

## Abstract

**Background:** Latino caregivers have poor mental health and access to caregiver support services. Here, we describe the protocol for a randomized controlled trial (RCT) to evaluate the effect of a text message intervention on depression and distress among informal Latino dementia caregivers. We will also assess mechanisms of action.

**Methods:** We are enrolling 288 Latino dementia informal caregivers 18 or older into a parallel group RCT. Participants randomized to the intervention group receive a remote, asynchronous, bilingual, bi-directional, six-month texting program focused on dementia education, skill-building and community resources. The control group enters a 7-month waitlist after which they are offered the same intervention as the intervention group. Randomization is stratified by each of the four recruitment sites at a 1:1 ratio. Outcomes (e.g., caregiver depressive symptomatology, distress) are measured via surveys at baseline, 3, 6 and 7 months.

**Conclusions:** This RCT addresse two priority areas: eliminating dementia disparities and optimizing caregiver support. Findings have the potential to make clinical and policy-relevant contributions by providing appropriate caregiver support to Latinos in a highly scalable way.

Trial registration: ClinicalTrials.gov #NCT06728436. University of Kansas Medical Center STUDY00150585.

## 1. Introduction

Family caregiving for people with Alzheimer’s disease and related dementias (ADRD) takes a serious mental health toll [1–9]. Latino family caregivers of people with ADRD experience substantial disparities and are often excluded from advances in research [10, 11]. The number of Latinos with ADRD is growing more than any other ethnic or racial group in the US [12]. Compared to non-Latino Whites, Latinos are more likely to become family caregivers [13], provide longer caregiving, and experience more caregiver distress and depression [8, 13–20]. Caregiver support interventions can improve mental health outcomes [21, 22]. However, most interventions have been designed for or tested among non-Latino Whites [10, 11], and evidence shows extant caregiver support interventions’ generalizability is questionable among Latinos [23–25]. Latinos experience unique barriers that might reduce traditional caregiver support interventions’ feasibility and efficacy, including transportation, financial, language, and cultural barriers [26, 27]. There is a critical need for efficacious caregiver support interventions for Latinos.

To address this need, we developed *CuidaTEXT* (Spanish for self-care and texting) using a set of user-centered principles [28]. *CuidaTEXT* is a multidomain, bilingual 6-month text message intervention that includes delivery and content features tailored to Latino caregivers. All caregivers receive one core daily automatic message on five domains: ADRD education, social support, self-care, ADRD care management, and behavioral symptoms. Caregivers can access on-demand automatic messages to expand on the five domains via sending keywords and interact with a coach via a live chat for more personalized help.

*CuidaTEXT* is the first ADRD caregiver support intervention to capitalize on text messaging. Newer and more fashionable technologies for caregivers exist such as websites, apps, and videocalls [29–31]. However, relying only on these newer technologies may perpetuate caregiver support disparities due to low access by Latinos [32]. *CuidaTEXT* was designed to be highly accessible among Latinos, as nearly 100% of Latino adults own a cellphone and 87% use texting (10% more than non-Latino Whites) [32, 33]. *CuidaTEXT’s* remote, asynchronous, and largely-automated features address important Latino caregiver barriers, including transportation, time constraints, and implementation costs of caregiver support interventions [34–37]. Its simple bilingual language and tailored content (e.g., resources for the uninsured, immigration-stress tips) address language, cultural, and contextual barriers.

Our one-arm feasibility study (n=24) showed *CuidaTEXT* was highly usable and acceptable, reduced caregiver stressors (e.g., behavioral symptom distress) and their negative consequences (e.g., depressive symptoms), and improved ameliorating factors of stress (e.g., approach-coping, preparedness for caregiving) at 6 months [28, 38]. In the current manuscript, we aim to describe the protocol for a randomized clinical trial (RCT) we are currently conducting to test the efficacy of *CuidaTEXT* among Latinos. To optimize intervention potency [39, 40], we will test mechanisms of change informed by the Stress Process Framework [41], the Science of Behavior Change, and our quantitative feasibility data [38].

## 2. Methods

### 2.1. Trial design and setting

This is a two-arm superiority RCT that uses parallel design with surveys administered at baseline, three, six months (end of treatment), and seven months (post-treatment; **Figure 1**). After completing the baseline assessment, the research team randomizes participants using a computer-generated randomization list implementing random permuted blocks that allocate participants to either CuidaTEXT or control at a ratio of 1:1, stratified by each of the four participating recruitment sites. Given the psychosocial nature of the study, intervention masking is only conducted at the data analyst level via color coding on REDCap, the data collection tool used in the project, so that they are not able to know which group participants were randomized to [42]. Enrollment started in February 2025 and is expected to end in September 2028. Four sites are conducting the recruitment of participants: One in Kansas (coordinating site), two in Florida, and one in Puerto Rico. All sites prescreen potential participants and refer candidates to the Kansas team, which completes the final screening, consents, conducts assessments, randomizes and provides coaching services within the CuidaTEXT intervention. Sites recruit from multiple sources including neurology and geriatric clinic visits, Electronic Health Records, community events, scientific and community presentations, social and ordinary media, research registries, community referrals, flyers, and word of mouth. An advisory board with caregivers, researchers, healthcare staff and community-serving professionals meets quarterly to guide recruitment efforts. For example, this advisory board has informed the addition of a texting option to hear more about the study, and simplify the project’s contact survey.

**Figure 1.**
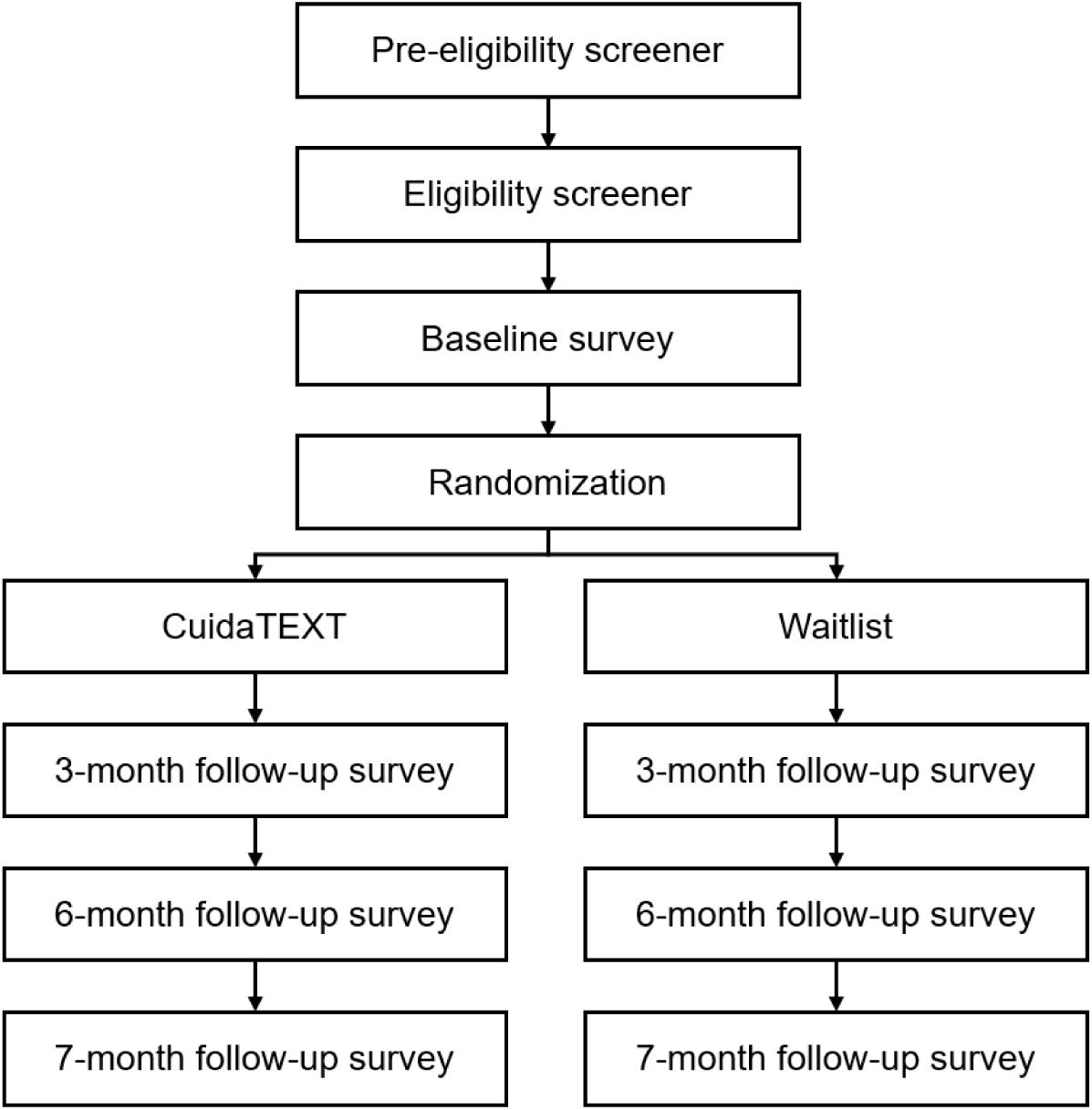
Flow diagram for recruitment and follow up in the CuidaTEXT Study.

Inclusion criteria includes Spanish and English-speaking people who are over the age of 18, identify as Latino, and report providing hands-on care for a relative who has been given a clinical or research ADRD diagnosis and has an Ascertaining Dementia-8 (AD-8) screening score ≥2 [43, 44], indicating cognitive impairment. Participants also need to have a score of 7 or higher on the 10-item Center for Epidemiologic Studies Depression Scale (CESD-10) [45] to avoid floor effects in our primary outcome (depressive symptomatology). In our feasibility study, effect sizes of improvements in depressive symptomatology increased as we limited the sample to those with higher baseline CESD-10 levels [38]. Participants are not eligible if they participate in another wellbeing-related clinical trial, cohabit with another participant, provide care for a person with ADRD already cared for by another CuidaTEXT participant, or plan to move to another country within seven months. Participants must self-report being able to read and write and use a cell phone at least weekly for texting. All types of cellphones are eligible. In the rare event that we encounter a potential participant who does not own a cell phone, we provide one for their use in the study which will be returned to the study team at the end of their participation.

All study procedures have been approved by the Institutional Review Board of the University of Kansas Medical Center (STUDY00150585), and recruitment sites rely of that IRB approval. All caregiver participants give written digital or in-person informed consent, unless when participants are unable to access and complete these consents, in which case their consent signature is waived, and a verbal consent is obtained and documented. This study was registered in ClinicalTrials.gov under the ID NCT06728436.

### 2.2. Eligibility, screening, consent, assessment, randomization, enrollment, and retention

The site recruitment teams provide potential participants with study information in their preferred language adapted for people with low literacy levels, either in person, at any of the recruitment sites, via phone, or videocall. After providing study information, they obtain a verbal consent and assess participants’ preliminary eligibility. Those preliminarily eligible for continuing screening are referred to the study operations team, which assigns the staff member who will complete the full eligibility assessment, signed consent, randomization, and baseline and follow-up assessments with that participant. Sites let potential participants know that they will receive a phone call from a Kansas phone number to reduce the risk of mistaking the call for spam. In line with methods to optimize retention [46], the team sets clear expectations for potential participants (e.g., length of assessments), explains the scientific principles behind trial methods (e.g., importance of retention irrespective of the group allocated to), helps diffuse ambivalence about participating (e.g., encourage to weigh pros and cons of participation). Randomized individuals are considered enrolled.

With regards to retention, study operations staff contact participants in both study arms to schedule an appointment two weeks before each assessment timepoint. They make multiple text, call, and email attempts as needed to schedule participants, and send reminder text messages two weeks before the timepoint and one day before the assessment day. Assessments are mostly remote but allow an in-person alternative as needed. During the eligibility screening, we collect participants’ and one to three relatives’ addresses, emails, and two telephone numbers to facilitate follow-up. Those who cannot be reached at follow-up are sent a certified letter to update the contact information. The team keeps flexible hours and allow assessments to be remote or in-person at the participants’ preferred location. We compensate participants with prepaid cards for baseline ($30), 3-month ($30), 6-month ($60), and 7-month assessments ($30). We waive the collection of social security numbers for compensation purposes to reduce the risk of stress.

### 2.3 Interventions

The intervention group receives *CuidaTEXT* immediately. This six-month intervention has been described earlier [28, 38]. CuidaTEXT entails sending 1-3 automated daily messages covering various aspects such as logistical instructions, ADRD education, social support, self-care, ADRD care management, and behavioral symptoms. Additionally, participants can text keyword-based queries for immediate assistance on the above-mentioned topics and engage in live chat sessions with the coach for further guidance upon request. Coaches’ responses are protocolized in a brief and simple manual to address these needs. For example, coaches will remind participants requesting more messages on a specific domain to use the keywords available as many times as necessary and visit specific websites or call the number of a national entity that can provide further help. Coaches will assist participants requesting help finding a specialist in their region using an online provider finder.

The intervention is guided by the Stress Process Framework [41, 47], which poses that caregivers’ background, context, and stressors have negative health consequences. Ameliorating factors of stress can reduce those stressors and its negative consequences. *CuidaTEXT’s* goal is to reduce stress resulting from caregiving and its negative consequences by targeting known ameliorating factors of stress. *CuidaTEXT’s* delivery and content features were designed to engage stress ameliorating factors (e.g., coping strategies and preparedness for caregiving) while addressing common Latino context, background, and stress-related barriers as follows:

#### Delivery features

These features were integrated to avoid adding stressors (e.g., commitment to in-person live interventions) and to reduce context and background barriers (e.g., transportation, time constraints).

- <u>Time constraints</u>. *CuidaTEXT* is asynchronous. Participants can use *CuidaTEXT* at any time. Caregivers receive only one message per day with core information to reduce burden. Caregivers can request additional on-demand messages as the program progresses based on their needs. Messages sent after office hours are either responded automatically if they are keywords or within the next two business days if these require a response from a coach.
- <u>Transportation barriers</u>. *CuidaTEXT* is remote. Participants can use the intervention anywhere.
- <u>Technological divide</u>. *CuidaTEXT* works via Short Message Service (SMS), which is integrated in all cellphones by default, is easy to use, and does not require a home broadband. Nearly 100% of Latino adults own a cellphone with texting capabilities [29–32]. Focusing solely on apps, computers, websites, or videocalls, which are underused by Latinos, could perpetuate caregiver support access disparities [29–32].
- <u>Financial difficulties</u>. Most of *CuidaTEXT* is automatic, relying little on workforce to reduce costs. Text message interventions for other conditions are more cost-effective than standard-of-care [34, 35].
- <u>Language barriers</u>. *CuidaTEXT* is available in English and US Spanish and does not use local slang or specialized jargon.
- <u>Literacy limitations</u>. *CuidaTEXT* was co-developed with the community to ensure text messages are easy to understand among Latinos [28]. During the development of the messages, we followed the Seven Principles of Communication: completeness, concreteness, courtesy, correctness, clarity, consideration, and conciseness [48].
- <u>Cultural aspects</u>. Receiving text messages on a private cellphone may address ADRD stigma [9, 36].

#### Content features

CuidaTEXT’s five domains described earlier target ameliorating factors of stress (e.g., approach-coping and preparedness for caregiving). Content considers key context and background factors as follows:

- <u>Cultural aspects</u>: All domains include testimonials from our Latino caregiver advisory board using mock Latino names. Video demonstrations for skill-building include Latino individuals. These content features address cultural congruence preferences [49, 50]. Tips for self-care or ADRD care management include cultural values (e.g., Latin dancing and healthy Latin American recipes). Tips for coping with behavioral symptoms address common Latino stressors, including the care recipient missing their home country or traveling to their home country for holidays. Given the centrality of family among Latinos [51], *CuidaTEXT* includes substantial content on family support and encourage forwarding text messages to their relatives.
- <u>Transportation, language, financial, and digital barriers</u>. Messages include links to community resources that are bilingual, remote, free, or that provide financial aid (e.g., remote caregiver support groups, respite grants, discounts for medications).

We ensure fidelity in delivering *CuidaTEXT* by integrating research process phases proposed by the Treatment Fidelity Workgroup of the NIH Behavior Change Consortium.[52] These include: 1) standardizing most of the intervention delivery via daily automatic and keyword-driven text messages, 2) standardizing coach protocols, 3) the coach supervisor completing a checklist weekly to track the history of the coach’s text message interactions (response times, legibility, and adequacy to caregiver needs), and providing feedback, 4) preventing contamination by excluding participants from the same household, and 5) planning for implementation setbacks by following up with participants to identify and address technical issues. To explore the impact of differential text message interaction, we track the frequency of all interactions between *CuidaTEXT* and participants and the time it takes the coach to respond to each live chat message. We will assess correlations between these frequency/time variables and change in outcomes.

Participants allocated to the control group are offered the opportunity to start *CuidaTEXT* after their 7 Month follow-up assessment, to increase participation interest. A waitlist control group serves as a benchmark to assess the effects of the intervention against not receiving treatment during that same time-period. This solution is more ethical than no treatment and may increase recruitment and retention, as all participants will ultimately receive *CuidaTEXT*. Waitlist control groups are common in caregiver support research, allowing comparability [21] and control for non-specific treatment effects [53]. Secondary analyses comparing *CuidaTEXT’s* efficacy at Month 7 (1-month post-treatment) will reduce the effects of attention associated with being in *CuidaTEXT,* which may drive self-reported improvements.

### 2.4 Assessments

This project has three specific aims informed by the Stress Process Framework: determine the effects of *CuidaTEXT* on Latino caregiver stressors and their negative consequences compared to a waitlist group (outcomes; Aim 1), explore CuidaTEXT’s effects on ameliorating factors of stress (targets; Aim 2) and examine the mediation of ameliorating factors on *CuidaTEXT’s* outcomes (Aim 3; Figure 2). Hypotheses include: Reductions in depressive symptoms (H1) and behavioral symptom distress (H2) will be higher in *CuidaTEXT* than control at Month 6 (Aim 1); Baseline to Month 6 increases in approach-coping (H3) and preparedness for caregiving (H4) will be higher in *CuidaTEXT* than control (Aim 2); and *CuidaTEXT’s* effects on change in depressive symptoms and behavioral symptom distress from Month 3 to 6 will be mediated by change in approach-coping (H5-H6) and preparedness for caregiving (H7-8) from baseline to Month 3 (Aim 3). All surveys include scales for outcomes and targets. See Table 1 for a schedule of assessments and interventions. Outcomes include:

**Figure 2.**
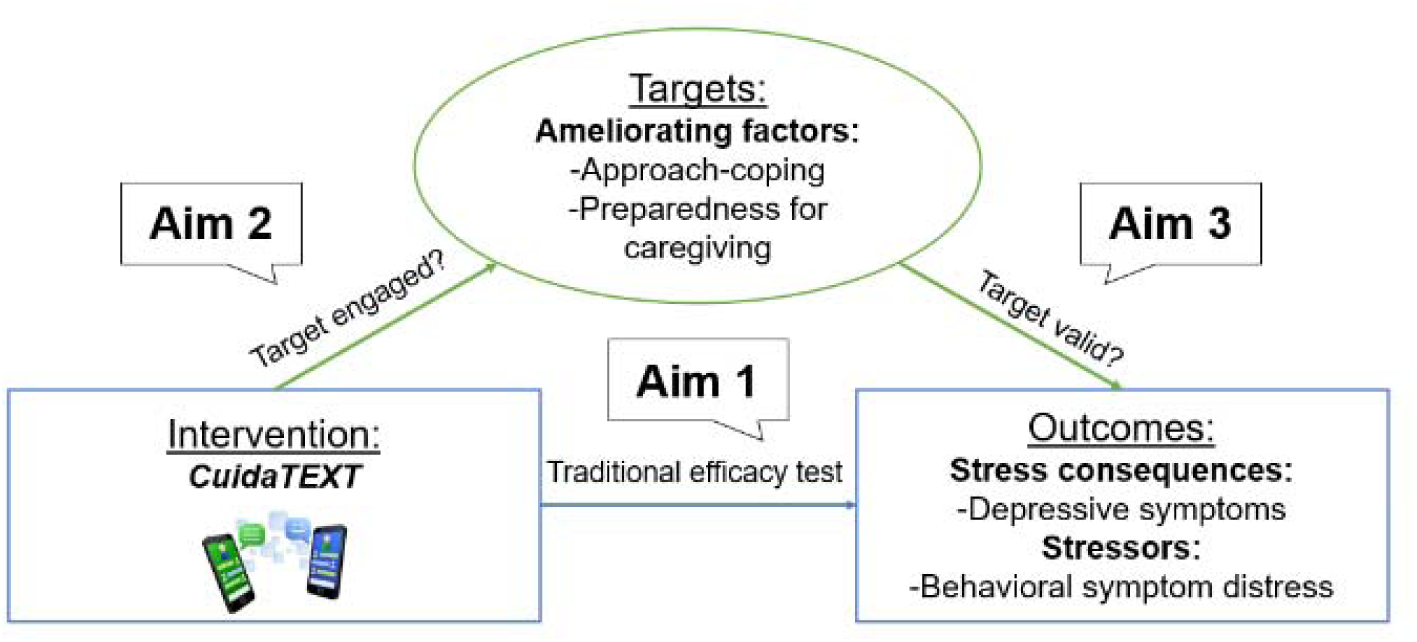
CuidaTEXT’s hypothesized mechanism of change.

**Table 1.**
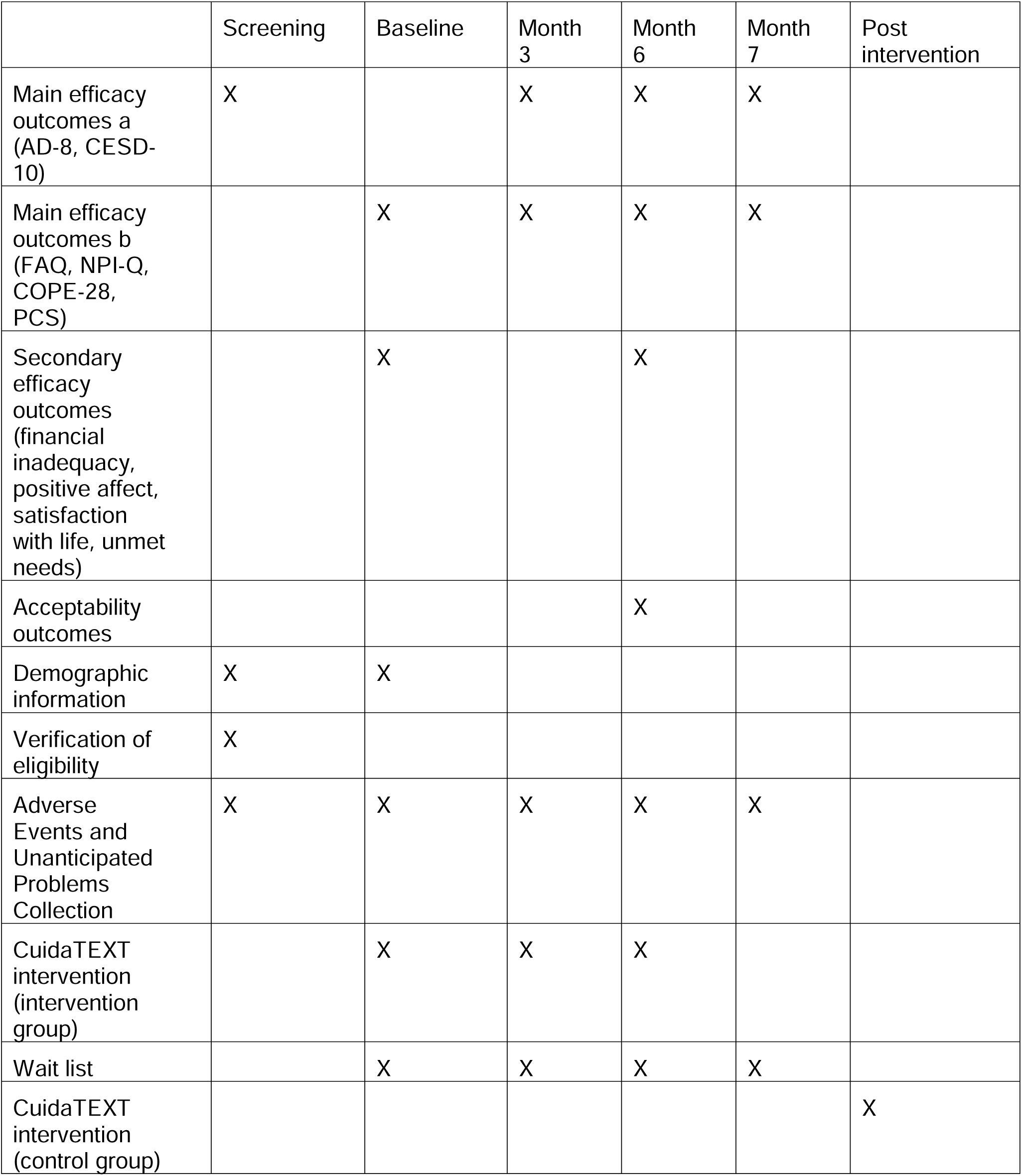
Timeline of assessments and interventions in the CuidaTEXT Study.

|  | Screening | Baseline | Month 3 | Month 6 | Month 7 | Post intervention |
| --- | --- | --- | --- | --- | --- | --- |
| Main efficacy outcomes a (AD-8, CESD-10) | X |  | X | X | X |  |
| Main efficacy outcomes b (FAQ, NPI-Q, COPE-28, PCS) |  | X | X | X | X |  |
| Secondary efficacy outcomes (financial inadequacy, positive affect, satisfaction with life, unmet needs) |  | X |  | X |  |  |
| Acceptability outcomes |  |  |  | X |  |  |
| Demographic information | X | X |  |  |  |  |
| Verification of eligibility | X |  |  |  |  |  |
| Adverse Events and Unanticipated Problems Collection | X | X | X | X | X |  |
| CuidaTEXT intervention (intervention group) |  | X | X | X |  |  |
| Wait list |  | X | X | X | X |  |
| CuidaTEXT intervention (control group) |  |  |  |  |  | X |

#### Caregiver depressive symptomatology

The primary outcome is the change in caregiver depressive symptomatology. We use the CESD-10 [54]. The CESD-10 is a self-report rating scale that measures characteristic symptoms of depression in the past week (e.g. depression, loneliness). Each item is rated on a 4-point scale (Rarely or None of the Time-Most or All of the Time). The summary score ranges from 0 to 30 with higher scores indicating higher depressive symptom severity. This scale has shown strong psychometric properties among English and Spanish-speaking Latinos [45, 55].

#### Behavioral symptom distress

The secondary outcome is the change in behavioral symptom distress. We use the Neuropsychiatric Inventory–Questionnaire (NPI-Q) [56, 57]. If any of 12 care recipient behavioral symptoms is present in the last month (e.g., repeating, delusions), caregivers rate their own distress on a 6-point scale (Not at All-Extreme). A distress summary score is calculated by adding the scores of all items. Higher scores indicate higher distress. The NPI-Q distress is responsive to change [58], and has shown strong psychometric properties among English and Spanish-speaking Latinos [55].

#### Targets include

#### Approach-coping

The primary target is change in approach-coping. We use the Coping Orientation to Problems Experienced Inventory (COPE-28) [59, 60]. The COPE-28 is a self-report questionnaire designed to measure ways to cope with caregiving. The scale is rated using a 4-point Likert scale (Not at All-a Lot). The approach-coping subscale uses 12 items corresponding with active coping, positive reframing, planning, acceptance, seeking emotional support, and seeking informational support [61]. High scores indicate caregiver participants use approach-coping techniques more often. The COPE-28 has an English [59] and a Spanish version [60], and the approach-coping subscale achieved an alpha coefficient of 0.8 in our feasibility study [38].

#### Preparedness for caregiving

The secondary target is change in preparedness for caregiving. We use the Preparedness for Caregiving Scale (PCS) [62, 63]. The PCS is a self-rated instrument that consists of eight items that ask caregivers how well prepared they believe they are for multiple domains of caregiving. Responses are rated on a 5-point Likert scale (Not at All Prepared-Very Well Prepared). The total score is the mean of all items and ranges from 0 to 4, with higher scores indicating higher preparedness. The scale has an English [62] and a Spanish version [63] and had an alpha coefficient of 0.8 in our feasibility study [38].

We also collect background, context and stressor variables, adverse events and unanticipated problems.

#### Background, context, and stressor variables

We collect background (age, sex, educational level, English proficiency, Latino subgroup [e.g., Mexican, Puerto Rican], ADRD subtype) and context variables (insurance status, rural setting, site) at baseline using English and Spanish items from reputable epidemiological surveys to assess background and context variables [64, 65]. We collect stressor variables at all timepoints using English and Spanish versions of the Functional Activities Questionnaire (FAQ; functional dependency) and the AD8 (cognitive status) [43, 44, 57, 66]. These variables are relevant for Latino health and ADRD research [67, 68].

#### Adverse events and Unanticipated problems

We collect AEs and Serious AEs (SAEs) in this trial and monitor for unanticipated problems through participant complaints. In line with successful caregiver support interventions, we have developed Standard Operating Procedures (SOPs) for tracking and reporting AEs, SAEs and unanticipated problems [69]. We have also developed SOPs for emergencies, suicidality, homicidality, and abuse. Our study staff is trained in these SOPs and notifies the PI of any suspicious information. The PI classifies reported events as AEs or unanticipated problems.

### 2.5. Sample size and data analysis

Aim 1 evaluates change in caregiver depressive symptomatology (CESD-10) from baseline to Month 6. In our feasibility study, caregivers with CESD-10 ≥7 had a mean 5.3-point reduction in CESD-10 scores (SD=5.2) [38], compared to <0.7-point reduction in controls in other studies [70–72]. A 3.5-point reduction (30%) is considered clinically meaningful [73], and thus our target effect. A sample of 114 caregivers per group will provide 99% power (α=0.05) to detect a difference between groups using a two-sided independent samples t-test. Assuming 25% attrition, we will recruit 144 per group. Secondary analyses will compare changes in caregiver distress (NPI-Q). Our feasibility data showed a 7.4-point mean reduction (SD=8.3), compared to 0.2 in controls [74]; this exceeds the 3.1-point minimal clinically important difference [75], yielding ≥80% power with 114 per group. Additional analyses will explore change in these outcomes at different timepoints (e.g., baseline to three or seven months).

Primary analyses will use intent-to-treat, imputing missing change scores (including death) as 0. We will examine missing data patterns and apply multiple imputation if needed. Completer-only and transformed variable analyses will be conducted as secondary checks.

Aim 2 compares CuidaTEXT vs control in changes in COPE-18 approach-coping and PCS preparedness scores using Bonferroni-corrected t-tests (α=0.025). Feasibility data showed 3.54-point (SD=6.79) and 0.53-point (SD=0.45) increases, respectively [38]. Compared to literature-reported changes [75, 76], our sample size will provide 80% power to detect an effect size of 0.41 for coping and 99% power for preparedness (effect size=1.04).

Aim 3 assesses whether increases in coping and preparedness mediate intervention effects on depressive symptomatology and distress via joint significance tests (Bonferroni corrected α=0.013) [77]. Four mediation tests will be conducted: (1) coping change from baseline to Month 3 as mediator between treatment and depressive symptoms change from Month 3 to 6; (2) coping change as mediator between treatment and distress change; (3) preparedness change as mediator between treatment and depression changes; and (4) preparedness change as mediator between treatment and distress change. Each test was powered at ≥80% based on effect sizes from our feasibility data and those used in Aim 2 [38]. If both treatment effects on the mediator and mediator effects on outcomes are significant, we will consider the target a potential mediator. Structural equation models will follow Diaz et al. (2011) [78], modeling causal pathways based on time-sequenced changes. Power for the final structural models will also be ≥80%, supporting their ability to detect meaningful mediated effects.

## 3. Summary

### 3.1. Overview of the CuidaTEXT study

This RCT examines the efficacy of the first SMS texting intervention for ADRD informal caregiver support in reducing depressive symptomatology and caregiver distress among Latinos. Individuals are recruited in four US sites. The intervention is based on the Stress Process Framework and was co-created with Latino ADRD caregivers. The primary outcome includes depressive symptomatology change from baseline to six months. Secondary outcomes include changes in caregiver distress, preparedness for caregiving, approach coping. Mediation analyses explore these four outcomes with preparedness for caregiving and approach coping as targets.

### 3.2. Novel features of CuidaTEXT

This study will be the first to test the efficacy of a text message ADRD caregiver support intervention in any population and among Latinos in particular. The innovation is not the text message modality itself but its application to address Latinos’ multiple barriers to caregiver support. Existing intervention modalities (i.e. face-to-face, web/app-based) have low reach among Latinos and older age groups [1, 79]. Our key innovation will be a new model that delivers scalable, wider reaching, and more impactful interventions to all Latino caregivers of people with ADRD. Indeed, *CuidaTEXT* can deliver caregiver support readily available to nearly 100% of Latino adults via SMS text messages. Most caregiver support interventions have been developed for and tested with non-Latino Whites [11]. *CuidaTEXT* tailors its delivery and content features to the Latino community [10, 79]. Most caregiver interventions are hard to implement in real-world settings [1, 79]. *CuidaTEXT*’s heavy reliance on automated text messages and the use of simple protocols for coaches who do not need to be experts in ADRD makes it an innovative and sustainable modality that will increase intervention fidelity and will reduce workforce costs.

### 3.3. Limitations

While rare, a person might care for multiple relatives with ADRD. In these cases, we will record the number of care recipients and ask about the one with the most severe symptoms in measures such as the NPI-Q. We considered allowing family caregiver clusters, as Latino family members tend to share decision-making [51, 80]. However, we will not allow this option given Latinos’ low willingness to participate in clusters in our feasibility study [38]. We do not include long-distance caregivers, who comprise 15% of caregivers [81].

Newer technologies such as apps might be more popular among Latinos in the future. However, their use might also fade and never reach the level of accessibility of SMS text messages. Neglecting text messaging and focusing solely on newer technologies, all of which are less accessible among Latinos than SMS texting, can perpetuate already-existing disparities among Latino caregivers [32]. This project’s experience will inform future interventions using popular newer technologies such as Facebook and WhatsApp [82], and language processing tools that may further reduce reliance on coaches.

## 4. Conclusions

This manuscript outlines a protocol for testing the efficacy of a text message intervention for informal Latino ADRD caregivers. This work is a continuation of more than five years of co-creation and feasibility testing of this SMS texting intervention with and for Latino caregivers of people with ADRD. Besides testing its efficacy, we will test mechanisms of action to inform how to optimize the intervention’s potency. This work is the first of its kind to capitalize on the high accessibility of text messaging among Latinos to deliver culturally-tailored ADRD caregiver support to this underserved and under-resourced population. Findings have the potential to make clinical and policy-relevant contributions by providing appropriate caregiver support to Latinos in a highly-scalable way [83].

## Data Availability

There are currently no data to share as this is a protocol.

## Declaration of generative AI and AI-assisted technologies in the writing process

During the preparation of this work the first author used Chat GPT to improve the readability and language of the manuscript and the cover letter. After using this tool, the authors reviewed and edited the content as needed and take full responsibility for the content of the published article.

## Acknowledgements

JPP thanks the national and local organizations that have partnered with him to conduct present and past research since 2015. The research team thanks research participants included in all stages of this research as well as anyone who has contributed directly and indirectly to this research. The ideas and opinions expressed herein are those of the authors alone, and endorsement by the authors’ institutions or the funding agency is not intended and should not be inferred.

## References

1. National Academies of Sciences & Medicine, Families caring for an aging America. 2016, National Academies Press.

2. Alzheimer’s Association, 2016 Alzheimer’s disease facts and figures. Alzheimer’s & Dementia, 2016. 12(4): p. 459–509.

3. Bertrand, R.M., L. Fredman, and J. Saczynski, Are all caregivers created equal? Stress in caregivers to adults with and without dementia. Journal of Aging and Health, 2006. 18(4): p. 534–551.

4. Friedman, E.M., et al., US prevalence and predictors of informal caregiving for dementia. Health Affairs, 2015. 34(10): p. 1637–1641.

5. Kasper, J.D., et al., The disproportionate impact of dementia on family and unpaid caregiving to older adults. Health Affairs, 2015. 34(10): p. 1642–1649.

6. Moon, H., et al., Predictors of discrepancy between care recipients with mild-to-moderate dementia and their caregivers on perceptions of the care recipients’ quality of life. American Journal of Alzheimer’s Disease & Other Dementias®, 2016. 31(6): p. 508–515.

7. Ory, M.G., et al., Prevalence and impact of caregiving: A detailed comparison between dementia and nondementia caregivers. The Gerontologist, 1999. 39(2): p. 177–186.

8. Pinquart, M. and S. Sörensen, Ethnic differences in stressors, resources, and psychological outcomes of family caregiving: A meta-analysis. The Gerontologist, 2005. 45(1): p. 90–106.

9. Alzheimer’s Association, 2021 Alzheimer’s disease facts and figures. Alzheimer’s & Dementia, 2021. 17(3).

10. Dessy, A., et al., Non-pharmacologic interventions for Hispanic caregivers of persons with dementia: systematic review and meta-analysis. Journal of Alzheimer’s Disease, 2022. 89(3): p. 769–788.

11. National Academies of Sciences, E., and Medicine, Meeting the challenge of caring for persons living with dementia and their care partners and caregivers: A way forward, T.N.A. Press, Editor. 2021: Washington, DC.

12. Wu, S., et al., Latinos & Alzheimer’s Disease: New numbers behind the crisis. Projection of the costs for U.S. Latinos living with Alzheimer’s Disease through 2060. 2016, USC Edward R. Roybal Institute on Aging and the LatinosAgainstAlzheimer’s Network.

13. National Alliance for Caregiving, Evercare® Study of Hispanic Caregiving in the U.S. 2008.

14. Napoles, A.M., et al., Reviews: developing culturally sensitive dementia caregiver interventions: are we there yet? American Journal of Alzheimer’s Disease & Other Dementias®, 2010. 25(5): p. 389–406.

15. Gallagher-Thompson, D., et al., Tailoring psychological interventions for ethnically diverse dementia caregivers. Clinical Psychology: Science and Practice, 2003. 10(4): p. 423–438.

16. Talamantes, M.A. and M.P. Aranda, Cultural competency in working with Latino family caregivers. 2004.

17. Garcia, E.M.O., Caregiving in the context of ethnicity: Hispanic caregiver wives of stroke patients. 2000.

18. Hinton, L., et al., Neuropsychiatric symptoms in Latino elders with dementia or cognitive impairment without dementia and factors that modify their association with caregiver depression. The Gerontologist, 2003. 43(5): p. 669–677.

19. Hinton, L., et al., Dementia neuropsychiatric symptom severity, help-seeking patterns, and family caregiver unmet needs in the Sacramento Area Latino Study on Aging (SALSA). Clinical gerontologist, 2006. 29(4): p. 1–15.

20. Liu, C., et al., Systematic review and meta-analysis of racial and ethnic differences in dementia caregivers’ well-being. The Gerontologist, 2021. 61(5): p. e228–e243.

21. Walter, E. and M. Pinquart, How effective are dementia caregiver interventions? An updated comprehensive meta-analysis. The Gerontologist, 2020. 60(8): p. e609–e619.

22. Butler, M., et al., Care interventions for people living with dementia and their caregivers. Comparative Effectiveness Review, A.f.H.R.a. Quality, Editor. 2020: Rockville, MD.

23. Pendergrass, A., et al., Dementia caregiver interventions: A systematic review of caregiver outcomes and instruments in randomized controlled trials. International Journal of Emergency Mental Health and Human Resilience, 2015. 17(2): p. 459–68.

24. Gitlin, L.N., et al., Translating evidence-based dementia caregiving interventions into practice: State-of-the-science and next steps. The Gerontologist, 2015. 55(2): p. 210–226.

25. Butler, M., et al., Care interventions for people living with dementia and their caregivers. Comparative Effectiveness Review, 2020(231).

26. Monahan, D.J., V.L. Greene, and P.D. Coleman, Caregiver support groups: Factors affecting use of services. Social Work, 1992. 37(3): p. 254–260.

27. Scharlach, A.E., et al., Racial and ethnic variations in caregiver service use. Journal of Aging and Health, 2008. 20(3): p. 326–346.

28. Perales-Puchalt, J., et al., A Text Messaging Intervention to Support Latinx Family Caregivers of Individuals With Dementia (CuidaTEXT): Development and Usability Study. JMIR aging, 2022. 5(2): p. e35625.

29. Grossman, M.R., D.K. Zak, and E.M. Zelinski, Mobile Apps for Caregivers of Older Adults: Quantitative Content Analysis. JMIR mHealth and uHealth, 2018. 6(7).

30. Waller, A., et al., Computer and telephone delivered interventions to support caregivers of people with dementia: a systematic review of research output and quality. BMC geriatrics, 2017. 17(1): p. 265.

31. Kajiyama, B., et al., Helping Hispanic dementia caregivers cope with stress using technology-based resources. Clinical gerontologist, 2018. 41(3): p. 209–216.

32. Pew Research Center. Mobile fact sheet. 2021 [cited 2021.

33. Duggan, M., Cell phone activities 2013. 2013, Pew Research Center.

34. Guerriero, C., et al., The cost-effectiveness of smoking cessation support delivered by mobile phone text messaging: Txt2stop. Eur J Health Econ, 2013. 14(5): p. 789–97.

35. Zurovac, D., et al., Costs and cost-effectiveness of a mobile phone text-message reminder programmes to improve health workers’ adherence to malaria guidelines in Kenya. PLoS One, 2012. 7(12): p. e52045.

36. Schilling, L., et al., Text messaging in healthcare research toolkit. Center for Research in Implementation Science and Prevention (CRISP), University of Colorado School of Medicine, 2013.

37. Hall, A.K., H. Cole-Lewis, and J.M. Bernhardt, Mobile text messaging for health: a systematic review of reviews. Annual review of public health, 2015. 36: p. 393–415.

38. Perales-Puchalt, J., et al., A text message intervention to support latino dementia family caregivers (CuidaTEXT): feasibility study. Clinical Gerontologist, 2022: p. 1–16.

39. Onken, L.S., et al., Reenvisioning clinical science: unifying the discipline to improve the public health. Clinical Psychological Science, 2014. 2(1): p. 22–34.

40. Nielsen, L., et al., The NIH Science of Behavior Change Program: Transforming the science through a focus on mechanisms of change. Behaviour research and therapy, 2018. 101: p. 3–11.

41. Pearlin, L.I., et al., Caregiving and the stress process: An overview of concepts and their measures. The Gerontologist, 1990. 30(5): p. 583–594.

42. Harris, P.A., et al., Research electronic data capture (REDCap)—a metadata-driven methodology and workflow process for providing translational research informatics support. Journal of biomedical informatics, 2009. 42(2): p. 377–381.

43. Galvin, J., et al., The AD8 A brief informant interview to detect dementia. Neurology, 2005. 65(4): p. 559–564.

44. Pardo, C.C., et al., Assessing the diagnostic accuracy (DA) of the Spanish version of the informant-based AD8 questionnaire. Neurología (English Edition), 2013. 28(2): p. 88–94.

45. González, P., et al., Measurement properties of the Center for Epidemiologic Studies Depression Scale (CES-D 10): Findings from HCHS/SOL. Psychological assessment, 2017. 29(4): p. 372.

46. Jake-Schoffman, D.E., et al., Methods-Motivational Interviewing approach for enhanced retention and attendance. American Journal of Preventive Medicine, 2021. 61(4): p. 606–617.

47. Sörensen, S., et al., Dementia care: mental health effects, intervention strategies, and clinical implications. The Lancet Neurology, 2006. 5(11): p. 961–973.

48. Sureka, B., P. Garg, and P.S. Khera, Seven C’s of Effective Communication. AJR. American Journal of Roentgenology, 2018. 210(5): p. W243–W243.

49. Sue, S., In search of cultural competence in psychotherapy and counseling. Am Psychol, 1998. 53(4): p. 440–8.

50. Apolinário-Hagen, J., et al., Investigating the Persuasive Effects of Testimonials on the Acceptance of Digital Stress Management Trainings Among University Students and Underlying Mechanisms: A Randomized Controlled Trial. Frontiers in psychology, 2021. 12.

51. Gallagher-Thompson, D., et al., Recruitment and retention of Latino dementia family caregivers in intervention research: Issues to face, lessons to learn. The Gerontologist, 2003. 43(1): p. 45–51.

52. Bellg, A.J., et al., Enhancing treatment fidelity in health behavior change studies: best practices and recommendations from the NIH Behavior Change Consortium. Health psychology, 2004. 23(5): p. 443.

53. Whitehead, W.E., Control groups appropriate for behavioral interventions. Gastroenterology, 2004. 126: p. S159–S163.

54. Cheng, S.T. and A.C. Chan, The center for epidemiologic studies depression scale in older Chinese: thresholds for long and short forms. International Journal of Geriatric Psychiatry: A journal of the psychiatry of late life and allied sciences, 2005. 20(5): p. 465–470.

55. Perales-Puchalt, J., et al., Validation of Wellbeing Scales Among Informal Caregivers of Latinos With Alzheimer’s Disease and Related Dementias. Hispanic Journal of Behavioral Sciences, 2025. 47(1): p. 27–48.

56. Kaufer, D.I., et al., Validation of the NPI-Q, a brief clinical form of the Neuropsychiatric Inventory. The Journal of neuropsychiatry and clinical neurosciences, 2000. 12(2): p. 233–239.

57. Acevedo, A., et al., The Spanish translation and adaptation of the uniform data set of the National Institute on Aging Alzheimer’s Disease Centers. Alzheimer disease and associated disorders, 2009. 23(2): p. 102.

58. Perales-Puchalt, J., et al., Effectiveness of “Reducing Disability in Alzheimer’s Disease” among dyads with moderate dementia. Journal of Applied Gerontology, 2021. 40(10): p. 1163–1171.

59. Carver, C.S., You want to measure coping but your protocol’too long: Consider the brief cope. International journal of behavioral medicine, 1997. 4(1): p. 92–100.

60. Perczek, R., et al., Coping, mood, and aspects of personality in Spanish translation and evidence of convergence with English versions. Journal of personality Assessment, 2000. 74(1): p. 63–87.

61. Eisenberg, S.A., et al., Avoidant coping moderates the association between anxiety and patient-rated physical functioning in heart failure patients. Journal of behavioral medicine, 2012. 35(3): p. 253–261.

62. Carter, J.H., et al., Living with a person who has Parkinson’s disease: the spouse’s perspective by stage of disease. Movement disorders: official journal of the Movement Disorder Society, 1998. 13(1): p. 20–28.

63. Gutierrez-Baena, B. and C. Romero-Grimaldi, Development and psychometric testing of the Spanish version of the Caregiver Preparedness Scale. Nursing Open, 2021. 8(3): p. 1183–1193.

64. Nelson, D., Reliability and validity of measures from the Behavioral Risk Factor Surveillance System (BRFSS). Sozial und Praventivmedizin 2001. 46(1): p. S3–42.

65. Beekly, D.L., et al., The National Alzheimer’s Coordinating Center (NACC) database: the uniform data set. Alzheimer Disease & Associated Disorders, 2007. 21(3): p. 249–258.

66. Pfeffer, R.I., et al., Measurement of functional activities in older adults in the community. Journal of gerontology, 1982. 37(3): p. 323–329.

67. Hilgeman, M.M., et al., Testing a theoretical model of the stress process in Alzheimer’s caregivers with race as a moderator. The Gerontologist, 2009. 49(2): p. 248–261.

68. Gallo, L.C., et al., The Hispanic community health study/study of Latinos sociocultural ancillary study: Sample, design, and procedures. Ethnicity & disease, 2014. 24(1): p. 77.

69. Czaja, S.J., et al., Data and safety monitoring in social behavioral intervention trials: the REACH II experience. Clinical trials, 2006. 3(2): p. 107–118.

70. Finkel, S., et al., E-care: a telecommunications technology intervention for family caregivers of dementia patients. The American journal of geriatric psychiatry, 2007. 15(5): p. 443–448.

71. Martindale-Adams, J., et al., A trial of dementia caregiver telephone support. Canadian Journal of Nursing Research Archive, 2013: p. 30–49.

72. Czaja, S.J., et al., A videophone psychosocial intervention for dementia caregivers. The American Journal of Geriatric Psychiatry, 2013. 21(11): p. 1071–1081.

73. Haase, I., M. Winkeler, and H. Imgart, [Anchor-based ascertaining of meaningful changes in depressive symptoms using the example of the German short form of the CES-D]. Neuropsychiatr, 2016. 30(2): p. 82–91.

74. Gavrilova, S.I., et al., Helping carers to care--the 10/66 dementia research group’s randomized control trial of a caregiver intervention in Russia. Int J Geriatr Psychiatry, 2009. 24(4): p. 347–54.

75. Gonzalez, E.W., et al., Enhancing resourcefulness to improve outcomes in family caregivers and persons with Alzheimer’s disease: A pilot randomized trial. International Journal of Alzheimer’s Disease, 2014.

76. Berry, A., T. Burke, and A. Carr, The impact of the first wave of the Covid-19 pandemic on parents of children with externalising difficulties in ireland: A longitudinal cohort study. Int J Clin Pract, 2021. 75(12): p. e14941.

77. Fritz, M.S. and D.P. MacKinnon, Required sample size to detect the mediated effect. Psychological science, 2007. 18(3): p. 233–239.

78. Diaz, F., et al., Using structural equations to test for a direct effect of some antipsychotics on triglyceride levels in drug-naïve first-episode psychosis patients. Schizophr Res, 2011. 131(1-3): p. 82–89.

79. Llanque, S.M. and M. Enriquez, Interventions for Hispanic caregivers of patients with dementia: A review of the literature. American Journal of Alzheimer’s Disease & Other Dementias®, 2012. 27(1): p. 23–32.

80. Apesoa-Varano, E.C., et al., Multi-cultural caregiving and caregiver interventions: A look back and a call for future action. Generations, 2015. 39(4): p. 39–48.

81. Cagle, J.G. and J.C. Munn, Long-distance caregiving: a systematic review of the literature. Journal of gerontological social work, 2012. 55(8): p. 682–707.

82. Adkins, D. and H.M. Sandy, Information seeking among latinos in the midwestern United States. Proceedings of the Association for Information Science and Technology, 2017. 54(1): p. 483–486.

83. Glasgow, R.E., et al., Evaluating the impact of health promotion programs: using the RE-AIM framework to form summary measures for decision making involving complex issues. Health education research, 2006. 21(5): p. 688–694.

